# Automated and semi-automated contact tracing: Protocol for a rapid review of available evidence and current challenges to inform the control of COVID-19

**DOI:** 10.1101/2020.04.14.20063636

**Authors:** Isobel Braithwaite, Tom Callender, Miriam Bullock, Robert W Aldridge

## Abstract

**Introduction:** Traditional approaches to case-finding, case isolation, and contact tracing methods have so far proved insufficient on their own to prevent the development of local epidemics of COVID-19 in many high-income countries despite relatively advanced public health systems. As a result, many governments have resorted to widespread social distancing measures and mass quarantines (‘lock-downs’) to reduce transmission and to prevent healthcare systems from being overwhelmed. However, such measures impose heavy human and societal costs. Automated or semi-automated digital contact tracing, in conjunction with scaled-up community testing, has been proposed as a key part of exit strategies from lockdowns. However, the effectiveness of these approaches to contact tracing is unclear, and to be effective, trusted, and widely adopted such technology must overcome several challenges.

**Methods and analysis:** We will perform a rapid systematic review to assess the effectiveness of automated and semi-automated digital tools for contact tracing, and identify key considerations for successful implementation, to inform the control of COVID-19. We will search PubMed, EMBASE, EBSCO Medical COVID information portal, OVID Global Health, Cochrane Library, medRxiv, BioRxiv, and arXiv for peer-reviewed and pre-print papers on automated or semi-automated digital tools for contact tracing of COVID-19, another respiratory disease with pandemic potential (limited to SARS, MERS, or pandemic influenza), or Ebola, in human populations. Studies will be eligible if published in English between 1 January 2000 and 14 April 2020. We will synthesise study findings narratively and will consider meta-analysis if ≥ 3 suitable studies with comparable interventions and outcomes are available.

**Ethics and dissemination:** Ethical approval is not required for this review. We plan to disseminate findings via pre-print, journal publication, through social media and web-based platforms and through direct stakeholder engagement.

## Introduction

COVID-19 is a highly infectious novel coronavirus whose spread has rapidly led to a global pandemic since the first cases were identified in China in late 2019 (Huang et al. 2020). In the absence of effective treatment or a vaccine, control of an emergent pathogen typically relies on effective case-finding (supported by testing), case isolation, and contact tracing, which is traditionally carried out manually using information provided by cases (Ferretti et al., 2020). Despite relatively advanced public health systems in many high-income countries, these methods have so far proved insufficient on their own to prevent the establishment of local epidemics, particularly without automation of contact tracing processes and where testing capacity is limited. To mitigate the impact of COVID-19 on health systems and reduce the number of deaths from the disease, many governments have resorted to measures including widespread social distancing, school and business closures, and mass quarantines (or ‘lock-downs’). However, these interventions impose heavy societal costs, including impacts on mental health, education, domestic violence and job losses; these are increased the longer lock-downs continue. Proposed exit strategies have emphasised the role of contact tracing in conjunction with substantially increased testing capacity (Ferretti et al., 2020).

Contact tracing is the process of finding individuals who are at increased risk of developing an infectious disease due to their exposure to a known case. This process is not conventionally automated, is time and resource intensive, and relies on cases’ accurate recall of information about their recent movements and interactions. Moreover, for it to have an impact case identification, diagnosis and contact tracing must all occur during the incubation period before a contact becomes infectious themselves. Because of this, contact tracing is rarely done for rapidly infectious respiratory infections, such as influenza. COVID-19 presents particular challenges for contact tracing (Hellewell et al. 2020): it is moderately transmissible, with the basic reproduction number (R_0_) estimated at 2-3 (Liu et al. 2020), its symptoms can be difficult to distinguish from other respiratory tract infections, it has a relatively short incubation period of approximately 5 days (Lauer et al. 2020), and a substantial proportion of individuals have asymptomatic clinical courses or mild symptoms that go relatively unnoticed (Bai et al. 2020). Moreover, a large majority of individuals are currently susceptible, such that large outbreaks of COVID-19 can develop rapidly.

To address challenges including imperfect recall by cases and the time delays often introduced by manual contact tracing (potentially enabling more complete, accurate and timely contact identification and notification), as well as to enable rapid scaling and reduce their resource-intensiveness, a number of novel automated or semi-automated approaches to contact tracing have been developed in the wake of the COVID-19 pandemic (e.g. Wang et al. 2020; [TraceTogether](https://www.tracetogether.gov.sg/); [Private Automated Contact Tracing](https://pact.mit.edu/) [PACT]). However, there are a number of potential pitfalls to such approaches, including concerns regarding privacy, security and other ethical considerations, and the fact that high levels of uptake by populations may be required for effectiveness (Ferretti et al. 2020; Hellewell et al. 2020). A review of the evidence regarding the effectiveness and implications of attempts to automate contact tracing to date may offer valuable learning to inform potential exit strategies from mass quarantines and technology development efforts, so that risks are addressed pre-emptively within any proposed strategy.

### Objective

To assess the effectiveness of automated and semi-automated digital tools for contact tracing, and identify key considerations for successful implementation, to inform the control of COVID-19.

## Methods and analysis

We will search PubMed, EMBASE, EBSCO Medical COVID information portal, OVID Global Health, Cochrane Library, medRxiv, BioRxiv, and arXiv, for pre-print and peer-reviewed articles from any geographical setting published from 1 January 2000 to 13 April 2020. Due to the short timelines for this review, our search is restricted to studies with a full text available in English. The search terms to be used for these databases are provided in Box 1. We will supplement this with a grey literature search using Google Advanced (with a sub-selection of these search terms due to Google Advanced’s restriction of searches to 32 search terms).

### Box 1: Search terms

Studies in which title, abstract or keywords (using MeSH terms where applicable) contain terms related to -

(“digital*” OR “automat*” OR “technology” OR “tech” OR “electronic*” OR “app” OR “application” OR “smartphone*” OR “smart-phone*” OR “mobile*” OR “phone*” OR “online*” OR “internet*” OR “m-Health” OR “mHealth” OR “e-Health” OR “eHealth” OR “telehealth*” OR “tele-health*”)

AND

(“contact trac*” OR “contact-trac*” OR “outbreak control” OR “outbreak prevention” OR “infectious disease control” OR communicable disease control” OR “health protection”)

AND

(“COVID*” OR “SARS-CoV-2” OR “novel coronavirus” OR “nCoV-2019” OR “MERS” OR “MERS-CoV” OR “SARS” OR “SARS-CoV” OR “H1N1” OR “swine flu” OR “H5N1” OR “avian flu” OR “avian influenza” OR “bird flu” OR “influenza*” OR “pandemic*” OR “ebola” OR “ebola hemorrhagic fever” OR “ebolavirus”)

Filters: English full-text only; 1 January 2000 to 13 April 2020 only

### Eligibility criteria

We will include the following study designs: interventional (randomised and non-randomised controlled studies), observational (cohort, case-control, cross-sectional, before-and-after studies, and time series analysis), modelling studies and case studies. Studies adopting solely qualitative study designs will not be included.

Articles must refer to automated or semi-automated digital tools for contact tracing in human populations of COVID-19, another respiratory disease with pandemic potential (limited to SARS, MERS, or influenza), or Ebola. We will restrict interventions to these disease groups to ensure that the results are applicable to COVID-19. Although Ebola is not a respiratory illness, novel methods for contact tracing in challenging circumstances were developed to contain this disease in conjunction with similar measures -for example, quarantines -to those applied in the current COVID-19 pandemic, such that findings may be applicable. The following digital tools will be eligible for inclusion: web-based (e.g. where a case inputs their contacts), stand-alone applications used on either a smartphone and/or computer, tools developed as an add-on to an existing application (e.g. adding a COVID-19 tracker to an existing health, mapping or messaging application), mobile signal traffic from telecommunications operators. We will include articles that provide a comparison either with no contact tracing, contact tracing without automated or semi-automated digital tools, other non-pharmaceutical disease control interventions, as well as articles without a comparator.

### Outcomes

Our primary outcomes of interest are the number or proportion of contacts identified and the number or proportion of contacts who go on to become cases who are identified. Secondary outcomes include: impact on R_0_ (or other indicators of outbreak control), uptake of the contact tracing tool or app, resource requirements or cost-effectiveness, data security and privacy, other ethical issues, public perception of the tool, lessons learnt from implementation of the intervention. Studies will be included in the review irrespective of whether primary or secondary outcomes of interest (or both) are reported. Studies written in languages other than English which appear relevant based on their abstracts will be reported in an appendix of possibly relevant articles.

### Study screening

Screening will be performed by two researchers with experience of public health and systematic review methods [IB and TC] using Covidence software. The initial (level one) screen of titles and abstracts will be carried out by one reviewer, with a second reviewer checking 10% of excluded records. Full-text (level two) screening of full manuscripts for eligibility will be undertaken by two reviewers, with one reviewer making initial inclusion decisions and a second reviewer checking all excluded records. A provisional screening form (available in the Appendix) has been prepared based on the eligibility criteria outlined above and has been pilot-tested by the review team using 10 citations for level one screening and 10 full-text articles for level two screening.

We will resolve any discrepancies by consensus between the two reviewers; a third reviewer [MB] will be involved in any cases where consensus cannot be reached through discussion. All decisions to exclude studies taken during both stages of study screening will be documented and outlined in the final report with a list of excluded studies.

### Data abstraction

Items for data abstraction will include:

- Study characteristics (e.g. study design, study setting (country), study period);
- disease(s) under study;
- Study population details (age range; other identifying characteristics);
- Selection method(s);
- Intervention details (device(s) tool is designed for -e.g. web browser, basic mobile phone, smartphone, multiple; automated or semi-automated; contact definition used / nature of contacts identified -e.g. face-to-face contact, proximity-based, other; technology used to identify recent contacts e.g. GPS, Bluetooth);
- Comparator details (a -no contact tracing; b -contact tracing without automated or semi-automated digital tools; c -other non-pharmaceutical disease control interventions);
- Primary outcomes (a -number or proportion of contacts (irrespective of subsequent case status) identified; b -number or proportion of contacts who go on to become cases who are identified);
- Details of any of the seven secondary outcomes of interest (see inclusion criteria) for which data is presented.

A standardized data abstraction form has been developed and pilot-tested to capture data on the above listed items.

### Data management

We will use the Covidence software platform for data management, and will report inclusion and exclusion of studies using a PRISMA flowchart. If there is a conflict between data reported across multiple sources for a single study (e.g. between a published article and a registry record), we will use data from the published article preferentially, taking into account any corrections or errata published subsequently.

### Quality assessment

For analytical studies including interventional and observational study designs, we will use the [Effective Public Health Practice Project (EPHPP)](https://merst.ca/ephpp/) tool (evaluated by Armijo-Olivo et al. 2012) and linked guidance to guide a rapid quality appraisal of studies eligible for inclusion. For economic evaluation and modelling studies we will use an adapted version of the [CHEERS checklist](https://www.equator-network.org/wp-content/uploads/2013/04/Revised-CHEERS-Checklist-Oct13.pdf) (Husereau et al. 2013), excluding any questions not relevant to the study design. Quality assessment will be undertaken by one reviewer for all included studies. Studies will not be excluded from narrative synthesis on the basis of the quality score assigned.

### Synthesis of findings

In addition to a summary table containing key details about each study, we will use narrative synthesis to bring together study findings. Meta-analysis will be considered if three or more papers investigating a comparable intervention within a similar disease context and which report a comparable quantitative primary outcome measure are identified.

### Ethics and dissemination

Ethical approval is not required for this review. We plan to disseminate findings via a pre-print server, journal publication, through social media and web-based platforms and through engagement with public health and policy stakeholders.

## Data Availability

Data availability is not relevant at this time because the submission is a rapid review protocol and no data exists. Extracted data in the final publication will be attached in an appendix and/or available upon request.

https://docs.google.com/spreadsheets/d/1Rj-Zh2JCZn2GoOqoP9iRU1yr9dU7h3CxfsEgSMX7Zoo/edit?usp=sharing

## Contributor statement

IB and RA conceived the study. All authors contributed to the development of the study. IB and TC drafted the protocol with critical input from RA and MB. IB and TC are guarantors of the review.

## Appendices

PRISMA-P form (template from Shamseer et al. 2015) completed and available here.

Study screening and data abstraction provisional template available [here](https://docs.google.com/spreadsheets/d/1Rj-Zh2JCZn2GoOqoP9iRU1yr9dU7h3CxfsEgSMX7Zoo/edit#gid=0).

## Full references

1. Armijo-Olivo S, Stiles CR, Hagen NA, et al. Assessment of study quality for systematic reviews: a comparison of the Cochrane Collaboration Risk of Bias Tool and the Effective Public Health Practice Project Quality Assessment Tool: methodological research. Journal of evaluation in clinical practice (2012): 18 (1); 12–18. doi: 10.1111/j.1365-2753.2010.01516.x

2. Bai Y, Yao L, Wei T, Tian F, Jin DY, Chen L, Wang M. Presumed asymptomatic carrier transmission of COVID-19. JAMA (2020). doi: 10.1001/jama.2020.2565

3. Ferretti L, Wymant C, Kendall M, et al. Quantifying SARS-CoV-2 transmission suggests epidemic control with digital contact tracing. Science (2020) doi: 10.1126/science.abb6936

4. Hellewell J, Abbott S, Gimma A, et al. Feasibility of controlling COVID-19 outbreaks by isolation of cases and contacts. Lancet Global Health. (2020) doi: 10.1016/S2214-109X(20)30074-7

5. Huang C, Wang Y, Li X, Ren L, Zhao J, Hu Y, Zhang L, Fan G, Xu J, Gu X, Cheng Z. Clinical features of patients infected with 2019 novel coronavirus in Wuhan, China. The Lancet (2020): 395 (10223); 497–506. doi: 10.1016/S0140-6736(20)30183-5

6. Husereau, D, Drummond M, Petrou S, et al. Consolidated health economic evaluation reporting standards (CHEERS) statement. International journal of technology assessment in health care (2013): 29(2); 117–122. doi: 10.1017/S0266462313000160

7. Lauer SA, Grantz KH, Bi Q, Jones FK, Zheng Q, Meredith HR, Azman AS, Reich NG, Lessler J. The incubation period of coronavirus disease 2019 (COVID-19) from publicly reported confirmed cases: estimation and application. Annals of internal medicine (2020). doi: 10.7326/M20-0504

8. Liu Y, Gayle AA, Wilder-Smith A and Rocklöv J. “The reproductive number of COVID-19 is higher compared to SARS coronavirus.” Journal of travel medicine (2020). doi: 10.1093/jtm/taaa021

9. Shamseer L, Moher D, Clarke M, Ghersi D, Liberati A, Petticrew M, Shekelle P, Stewart LA. Preferred reporting items for systematic review and meta-analysis protocols (PRISMA-P) 2015: elaboration and explanation. BMJ 349 (2015). doi: 10.1136/bmj.i4086

10. Wang, CJ, Ng CY, and Brook RH. Response to COVID-19 in Taiwan: big data analytics, new technology, and proactive testing. JAMA (2020) doi: 10.1001/jama.2020.3151

